# Pilot Randomised Controlled Trial - Nurture Early for Optimal Nutrition (NEON) Study: Community facilitator-led participatory learning and action (PLA) women’s groups to improve infant feeding, care and dental hygiene practices in South Asian infants aged < 2 years in East London

**DOI:** 10.1101/2024.03.07.24303745

**Authors:** Logan Manikam, Priyanka Patil, Tala El Khatib, Subarna Chakraborty, Delaney Douglas- Hiley, Sumire Fujita, Joanna Dwardzweska, Oyinlola Oyebode, Clare H. Llewellyn, Kelley Webb-Martin, Carol Irish, Mfon Archibong, Jenny Gilmour, Phoebe Kalungi, Neha Batura, Kalpita Shringarpure, Monica Lakhanpaul, Michelle Heys, NEON Steering Team Membership of the NEON Steering Team is listed in the Acknowledgements

## Abstract

**Background:** The first 1000-days of life are a critical window and can result in adverse-health consequences due to inadequate nutrition. South-Asian (SA) communities face significant health-disparities, particularly in maternal and child-health. Community-based-interventions, often employing Participatory-Learning-and-Action (PLA) approaches, have effectively addressed health-inequalities in lower-income-nations. The aim of this study was to assess the feasibility of implementing a PLA-intervention to improve infant-feeding and care-practices in SA communities in London.

**Methods:** Comprehensive-analyses were conducted to assess the feasibility/fidelity of this pilot-randomised-controlled-trial. Summary-statistics were computed to compare key-metrics (participant consent-rates, attendance, retention, intervention-support, perceived-effectiveness) against predefined-progression-rules guiding towards a definitive-trial. Secondary-outcomes were analysed, drawing insights from sources, such as The-Children’s-Eating-Behaviour-Questionnaire (CEBQ), Parental-Feeding-Style-Questionnaires (PFSQ), 4-Day-Food-diary, and the Equality-Impact-Assessment (EIA) tool. Video-analysis of children’s mealtime behaviour trends was conducted. Feedback-interviews were collected from participants.

**Results:** Process-outcome measures met predefined-progression-rules for a definitive-trial which deemed the intervention as feasible. The secondary-outcomes analysis revealed no significant changes in children’s BMI z-scores. This could be attributed to the abbreviated follow-up period of 6-months, reduced from 12-months, due to COVID-19-related delays. CEBQ analysis showed increased food-responsiveness, along with decreased emotional-over/undereating. A similar trend was observed in PFSQ. The EIA-tool found no potential discrimination areas, and video-analysis revealed a decrease in force-feeding-practices. Participant-feedbacks revealed improved awareness and knowledge-sharing.

**Conclusion:** The study validates the feasibility of a community-oriented, co-adapted Participatory-Learning-and-Action approach for optimising infant-care among South-Asians in high-income countries. It underscores the potential of such interventions in promoting health-equity and improving health-outcomes. Further research is required to evaluate their wider impact.

## Background

During the initial 1000 days of a child’s life, lifestyle decisions made during pregnancy and infancy significantly influence future health and cognitive outcomes, with poor nutrition in this period linked to adverse health effects such as diabetes, heart disease, and dental disorders. Conversely, optimal nutrition during this timeframe contributes to academic success and fewer behavioural problems^1,3^.

Ethnic minorities in the United Kingdom (UK), particularly South Asian (SA) communities, face significant disparities in maternal and child health^5^. Research on complementary feeding practices (CFP) among SA families in the UK reveals persistent non-recommended practices, despite adherence to WHO guidelines. Bicultural challenges, low acculturation levels, conflicting guidance from health professionals, and community leaders contribute to these practices. Barriers such as short birth intervals, poverty, and the perception of better uses of the mother’s time also hinder engagement in WHO-recommended CFP.

Traditionally, community-based interventions utilizing Participatory Learning and Action (PLA) have effectively addressed health disparities in low-income countries (LMICs). PLA emphasizes the efficacy of community-based interventions, highlighting their role in improving health outcomes and addressing social determinants among marginalized populations^1,11^. These interventions engage community members in program development, fostering health equity by addressing root causes like poverty and educational deficits, and aligning with community needs and cultural values^12,1^. Recent studies also highlight the effectiveness of community-based interventions in high-income countries (HICs), addressing issues like obesity, cardiovascular disease, mental health, and substance abuse^1^.

Community-based interventions involving PLA approaches, characterized by group discussions, role-playing, and problem-solving exercises, promote active learning, collaboration, and trust-building^1^. Led by community facilitators, the PLA approach follows a four-stage cycle, aligning with WHO-endorsed strategies to enhance maternal and infant survival in low-income countries^1, 18^. This approach has demonstrated effectiveness in reducing maternal and neonatal mortality, improving breastfeeding rates, and addressing health disparities ^1,1^.

To address knowledge gaps in infant feeding practices among UK ethnic minorities, we adapted and extended the successful community-based PLA approach to engage South Asian families in East London through the NEON program^2,21^.

NEON 1’s success-a community-based participatory study identified infant feeding practices and socioecological factors within the British-Bangladeshi community in East London through culturally tailored parenting interventions to optimise infants’ nutrition leading to NEON 2-focusing on evaluating the feasibility, fidelity, and acceptability of an online delivery option alongside the traditional face-to-face approach optimizing infant feeding, care, and dental hygiene practices^22, 23^. The pilot study seeks to evaluate the feasibility of advancing to a definitive trial, considering predefined criteria such as participant recruitment and retention rates, intervention support, and overall acceptability. Secondary objectives involve assessing data collection outcomes, blinding effectiveness, intervention delivery time requirements, and the primary outcome—child BMI.

## Data and Methods

### Study Design

A pilot feasibility single-blinded cluster Randomized Controlled Trial (RCT) with three arms was conducted in East London boroughs Tower Hamlets (TH) and Newham (NH). See supplemental material 1 for the trial design. Six high-density wards in each borough, outlined in the official protocol²², were selected for the study, detailing allocation, randomization, inclusion/exclusion criteria, outcome measures, and data analysis.

#### Sample

Initially designed for three boroughs (NH, TH, and Waltham Forest [WF]), the study planned 20 Women’s PLA groups (equally distributed in two intervention arms) and 32 Women’s PLA group cycles (16 in each intervention arm). Recruitment targeted 288 to 384 participants, with WF withdrawing due to COVID-19 concerns. Sample size adjustment was deemed unnecessary as TH and NH achieved recruitment objectives.

#### Patient and Public Involvement and Engagement

In this co-designed study, five Community Facilitators (CFs) from NEON 1 were promoted to Community Researchers (CRs) to aid in all aspects of intervention development and evaluation. Recruited CFs served as key collaborators, enhancing community engagement, and co-developing relevant topic guides. For NEON 2, an additional ten CFs were recruited and trained for intervention delivery. CRs contributed to the development of topic guides, questionnaires, data analysis, and dissemination activities.

### Intervention

The study involved eight biweekly sessions conducted over a period of 14 weeks. The sessions were divided into four phases - learn, identify, implement, and evaluate. The sessions were conducted face-to-face and online, as appropriate. Participants in the usual care arm received conventional standard care visits and did not participate in the Participatory Learning and Action (PLA) group sessions. Facilitators maintained contact with them for data collection.

#### Monitoring and Evaluation

The Research Assistant (RA) monitored facilitator feedback and met regularly with CFs to ensure consistency and structure. Based on discussions with CFs and CRs, the RA addressed issues by coaching and motivating facilitators towards study goals.

### Data Analysis

#### Summary of Baseline Data and Sample Characteristics

Demographic characteristics were summarized with means and standard deviations for continuous variables and percentages for categorical variables. Descriptive statistics covered quantitative data from participants’ demographic questionnaire, PLA cycle meeting register, direct observation checklists, and sustainability assessment.

#### Process Outcome Analysis

Quantitative data on recruitment and retention rates across the three trial arms were descriptively summarized using frequency, percentages, and 95% confidence intervals (CI). The screening, eligibility, consent, and randomization/registration processes were also summarized. Participant breakdown by ethnic/language group in each borough was provided. To evaluate compliance and fidelity in delivering NEON, the number of NEON Women’s Group PLA Cycles and attendance rates per arm were assessed and compared against predefined Stop/Go criteria (Table 2).

#### Trial Outcome Analysis

ANOVA was utilized to compare mean BMI z-scores across face-to-face, online, and control arms, with differences assessed from baseline to post-PLA sessions and six months post-intervention using STATA 17.0 with significance set at 0.05.

Children’s-Eating-Behaviour Questionnaire (CEBQ), Parental Feeding-Style Questionnaires (PFSQ), 4 Day Food-diary, and the Equality Impact-Assessment (EIA) tool were all administered via REDCap (a web-based application), yielding summary statistics and participant insights. Infant feeding videos were analysed using ELAN, with behaviours assigned to pre-established codes.

#### Sample Size and Power Calculations

To inform future trial sample size, one-way ANOVA evaluated inter-group and intra-group variance. G*Power Software computed the sample size, considering PLA BMI z-scores at study end and ANOVA insights. Further methodological details are in the published study protocol^²²^.

## Results

### Demographic data

Mothers’ ages ranged from 20 to 58, averaging 32 years (N = 241; SD 5.98) (Table 3). Children were 4 to 24 months old, averaging 12.4 months (N = 155; SD 5.19), with 45.1% males and 54.9% females (N = 133). The majority resided in NH (78.1%), and the rest in TH. Main mother ethnicities were Bangladeshi (65.4%), Pakistani (16.9%), Sri Lankan (7.4%), Indian Punjabi (6.9%), and Indian Gujrati (3.5%). Most children shared similar ethnicities, except for 4.21% being bi-racial.

A significant number of mothers were married (96.1%). Assistance in child care was reported by 65.5%, and 34.5% had no assistance. Regarding education, 40.5% were high school graduates, 12.1% had associate degrees, 28.8% had higher degrees, and 18.7% had less than a high school degree or no degree. Employment-wise, 70.0% engaged in homemaking, 10.1% worked part-time, and 19.8% were employed full-time. English literacy levels were self-rated as low or non-existent for 24.5%, moderate for 30.7%, and high (fluent) for 44.7%.

In terms of housing, 74.7% rented, 17.8% owned or had a mortgage, 6.2% lived rent-free, and 1.7% were in temporary accommodations. Annual family income varied, with 55.6% choosing not to disclose. The disclosed incomes ranged from £0 to £31,495 or more, with varying percentages for each income bracket.

**Table 1.** Participants’ Characteristics.

|  | **Categories** | **N (%)** |
| --- | --- | --- |
| **Mother's Age (Years)** | Not Applicable | Mean: 32 years<br>SD: 5.98 |
| **Infant's Age (Months)** | Not Applicable | Mean: 12.4 months<br>SD: 5.19 |
| **Infant's Sex** | Male<br>Female | 60 (45.1%)<br>73 (54.9%) |
| **Borough of Residence** | Newham<br>Tower Hamlets | 200 (78.1%)<br>56 (21.9%) |
| **Mother's Ethnicity** | Bangladeshi<br>Pakistani<br>Sri Lankan/ Tamil<br>Indian Punjabi<br>Indian Gujarati | 151 (65.4%)<br>39 (16.9%)<br>17 (7.4%)<br>16 (6.9%)<br>8 (3.5%) |
| **Infant's Ethnicity** | Bangladeshi<br>Pakistani<br>Sri Lankan<br>Indian Punjabi<br>Indian Gujarati | 137 (64.0%)<br>37 (17.3%)<br>13 (6.1%)<br>13 (6.1%)<br>14 (6.5%) |
| **Mother's Marital Status** | Single<br>Married<br>Widowed<br>Divorced | 8 (3.1%)<br>247 (96.1%)<br>1 (0.4%)<br>1 (0.4%) |
| **Mother's Highest Education Level** | Less than Secondary Education<br>Secondary Education<br>Associate Degree<br>Tertiary Education | 48 (18.7%)<br>104 (40.5%)<br>31 (12.1%)<br>74 (28.8%) |
| **Mother's Employment Status** | Unemployed/sick/retired<br>Part-time employee<br>Full Time employee | 173 (70.0%)<br>25 (10.1%)<br>49 (19.8%) |
| **Mother's English Literacy Level** | Little to None<br>Moderate<br>Fluent | 63 (24.5%)<br>79 (30.7%)<br>115 (44.7%) |
| **Housing** | Rent<br>Own of Mortgage<br>Temporary Accommodation<br>Rent-Free | 180 (74.4%)<br>43 (17.8%)<br>4 (1.7%)<br>15 (6.2%) |
| **Average Family Income** | £0-£5,475<br>£5,476 - £12,097<br>£12,098 - £20,75<br>£20,754 - £31,494<br>£31,495 or more<br>Did not disclose | 34 (14.7%)<br>25 (10.8%)<br>15 (6.5%)<br>16 (6.9%)<br>15 (6.5%)<br>129 (55.6%) |

### Process Outcomes

#### Screening, Eligibility, and Registration

Of the 263 individuals enrolled based on inclusion/exclusion criteria, only 186 consented, marking a significant decline. Nevertheless, with 71% of eligible participants consenting, surpassing the >=50% benchmark, the trial proceeded. Figure 1 provides a CONSORT diagram detailing enrolment, allocation, and attrition rates.

**Figure 1.**
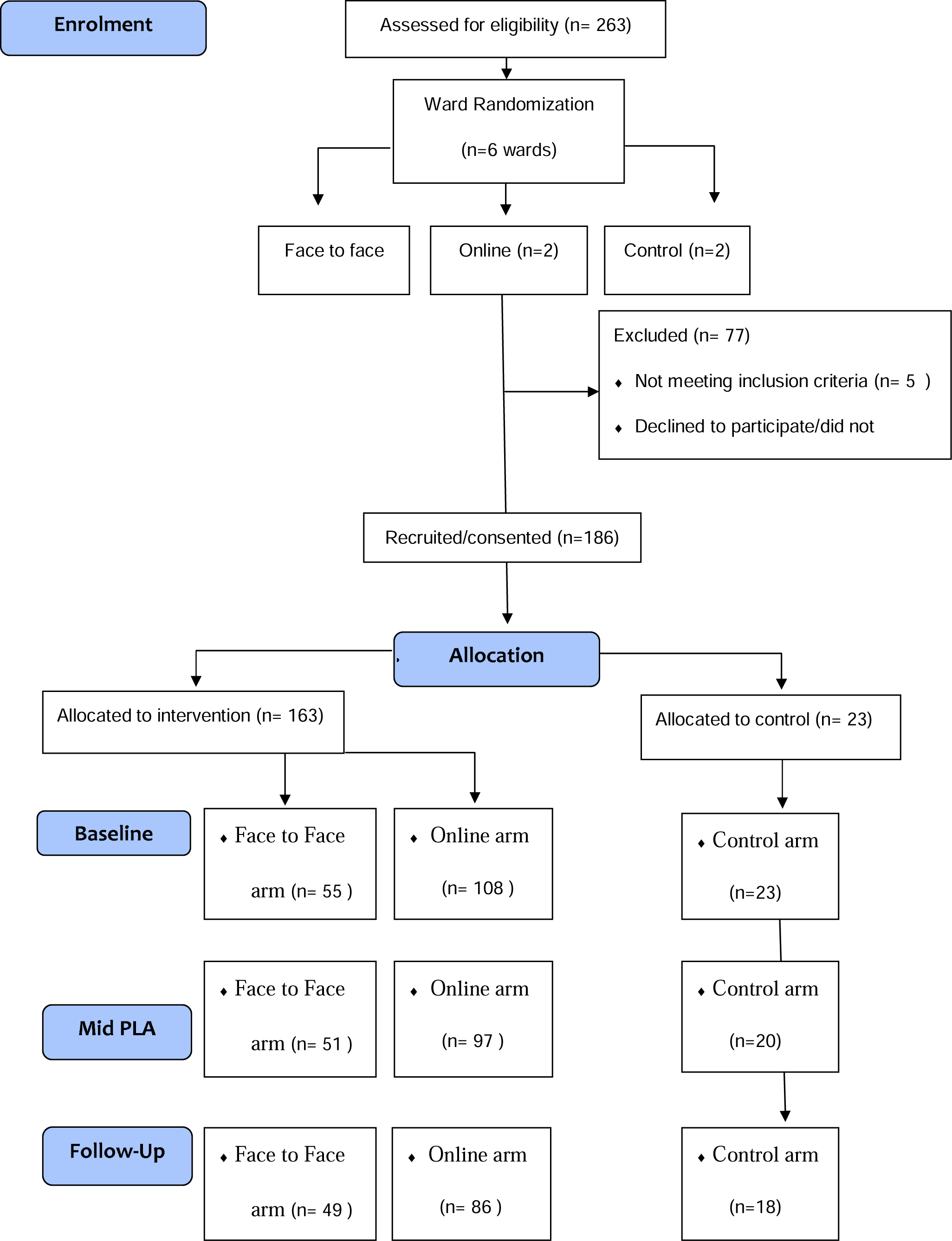
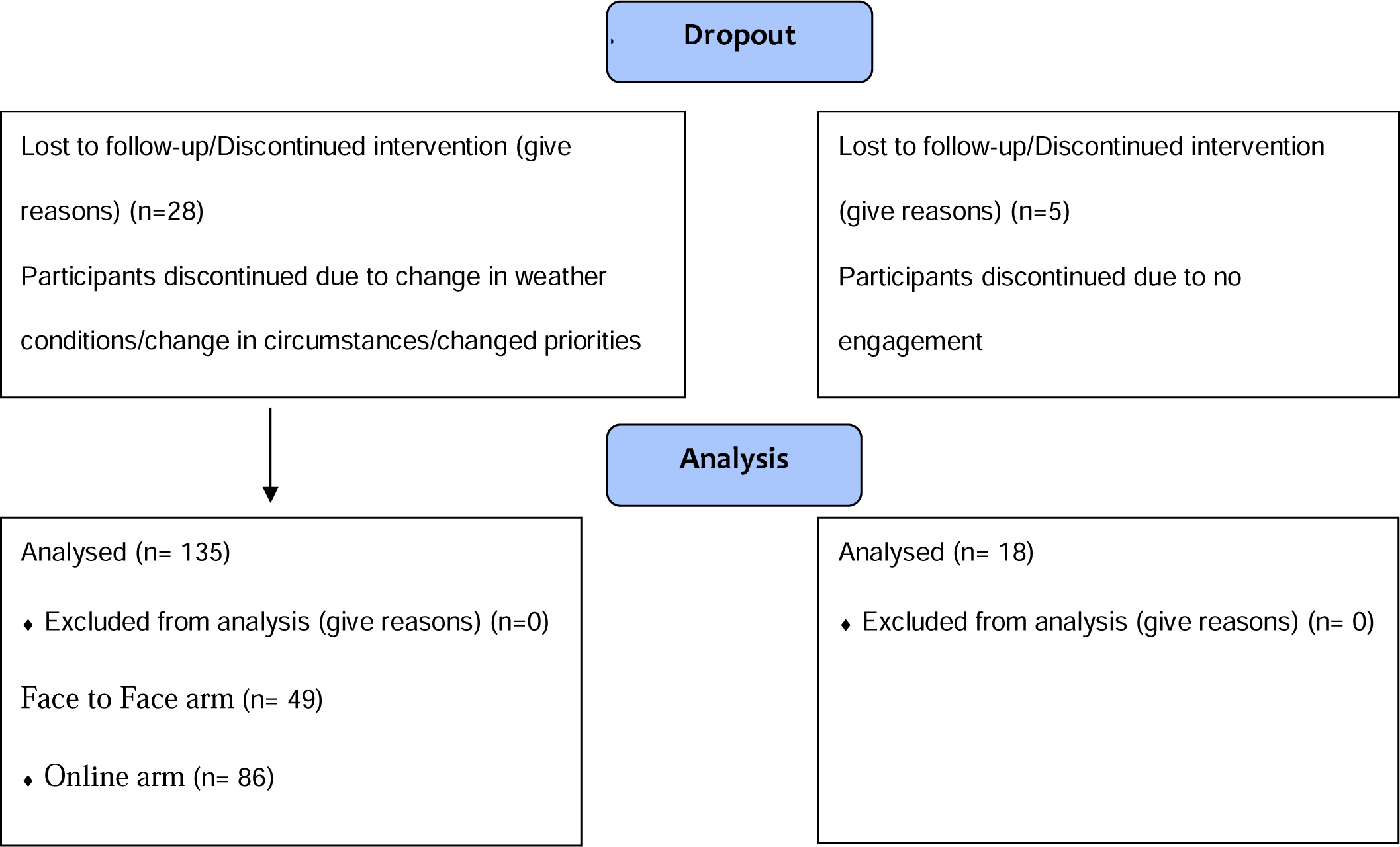
NEON RCT Consort Flow Diagram

NH had participants from Pakistani (13.9%), Punjabi (9.6%), Gujarati (11.3%), Tamil (8.7%), and Bangladeshi (56.5%) backgrounds. TH included 37 Bangladeshi participants. Refer to supplemental material 2 for the participant counts and ethnic breakdowns.

#### Sessions Planned and Delivered

The PLA sessions, initially comprising 25 groups (20 NH, 5 TH), experienced a reduction in numbers by the third cycle due to a participant drop from 186 to 167. This led to a decrease in sessions to 11 in NH and 4 in TH. By the fifth meeting, further reductions resulted in 4 face-to-face and 6 online sessions in NH, and 2 of each in TH. Concurrently, usual care participants decreased in NH, while remaining constant in TH.

#### Participant Attendance

In NH, the combined attendance rate for both intervention arms was 29%, while in TH, the attendance rate reached 59%. The overall attendance rate across all intervention arms was 37%, meeting the predefined “Go” criterion. For detailed attendance rates for PLA sessions, refer to supplemental material 3.

#### Retention Rates

The overall trial retention rate was 65%. In NH, retention rates were 50% face-to-face (29 of 59), 67% online (69 of 103), and 34% in the control arm, resulting in a total retention rate of 48%. In TH, retention rates were 71% in face-to-face (20 of 28), 88% in online (16 of 18), and 33% in the control arm, yielding a total retention rate of 78%. Height and weight measurements were completed by 65% of total participants, with TH at 79% and NH at 67% (29/39 in TH, 87/128 in NH).

#### Intervention Support

##### Participants’ Feedback

The questionnaire was distributed digitally via email, and due to a lack of responses, phone and face-to-face interviews were conducted with willing participants, garnering feedback from 46 individuals. Participants in the face-to-face arm appreciated in-person meetings, providing the chance to meet other mothers. However, they suggested an option for online attendance when face-to-face was impractical. The 2-hour session duration was deemed too lengthy for most participants. Sessions were conducted in native languages, with participants expressing a desire for translated toolkit content in future sessions.

##### Facilitators’ Feedback

A thematic analysis was conducted on participant and facilitator feedback to identify recurring patterns and comments. The objective was to explore their perspectives on intervention delivery and optimize the design, provision, and reach of a future definitive trial. Detailed results of the thematic analysis are summarized in a separate article.

##### Direct Observation of Intervention Delivery

CRs ensured PLA cycle consistency in women’s groups. Facilitator adherence was digitally assessed by independent observers using a 1-4 scale, with perceptions captured via open-ended questions (refer to supplemental material 4). Non-attendance led to undelivered sessions. Despite schedule changes, Bangladeshi CFs covered most delivery phase components. Challenges such as late arrivals and low attendance shortened sessions to 15-30 minutes. High-attendance sessions received positive feedback and fostered discussions. CFs demonstrated adaptability, modifying sessions in response to challenges, notably in PLAs 3 and 4. Online sessions prioritized crucial content, addressing some issues. However, time constraints prevented the implementation of solutions proposed in PLA 7. PLA 8, deviating from its primary objective of evaluating implemented solutions, scored the lowest.

##### Sustainability Assessment

CRs employed a sustainability assessment tool to self-assess group capacity across nine key domains for sustainability and community empowerment. The Bangladeshi group consistently scored highest in every domain. Figure 2 illustrates diverse strengths and weaknesses across groups. Notably, participation scores appear to influence other domains, with higher participation correlating with increased mean scores and higher ratings in other components. For a summary of process outcomes against pre-established Go/Stop criteria, see Table 2.

**Figure 2.**
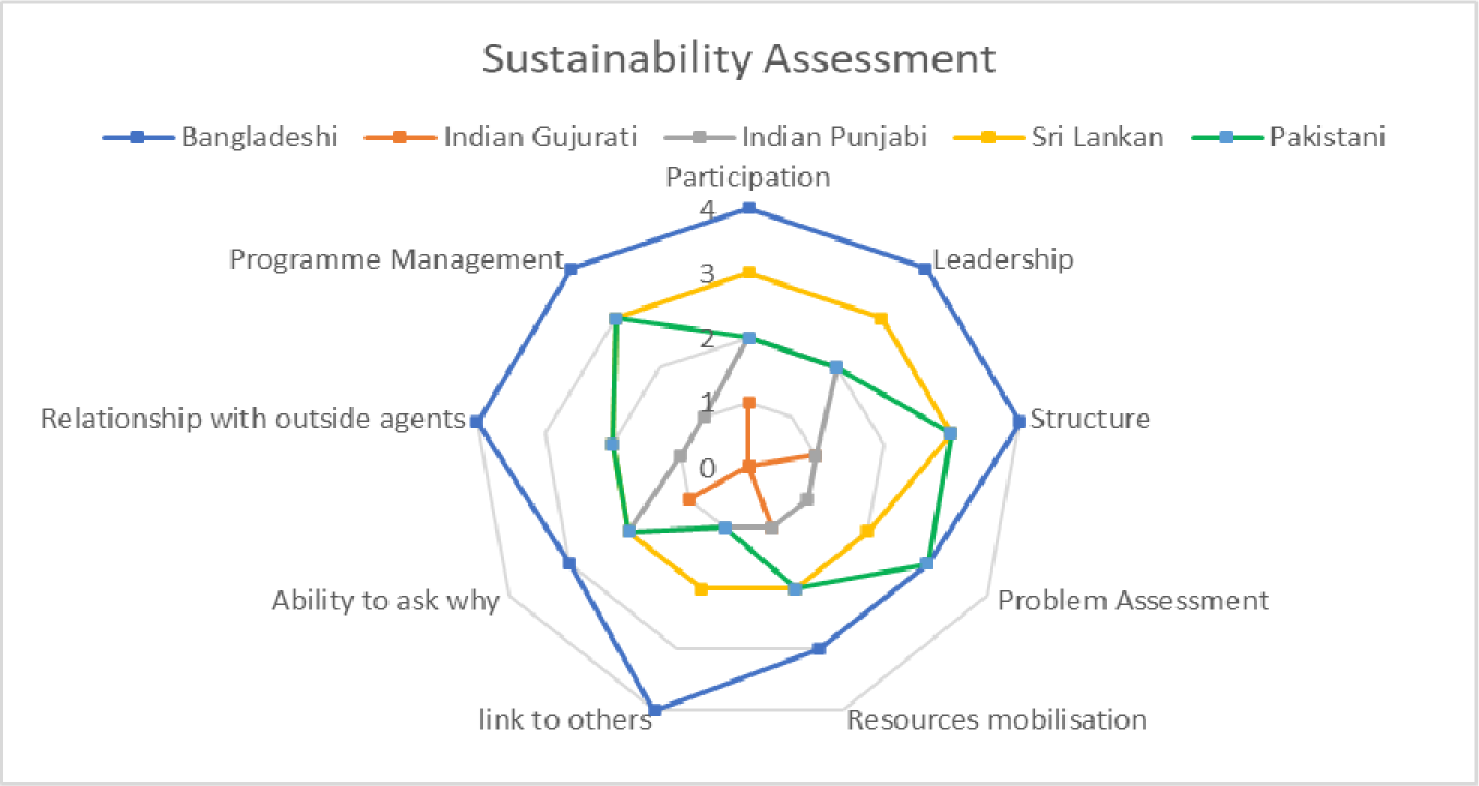
Visual representation of sustainable assessment scores

**Figure 3.**
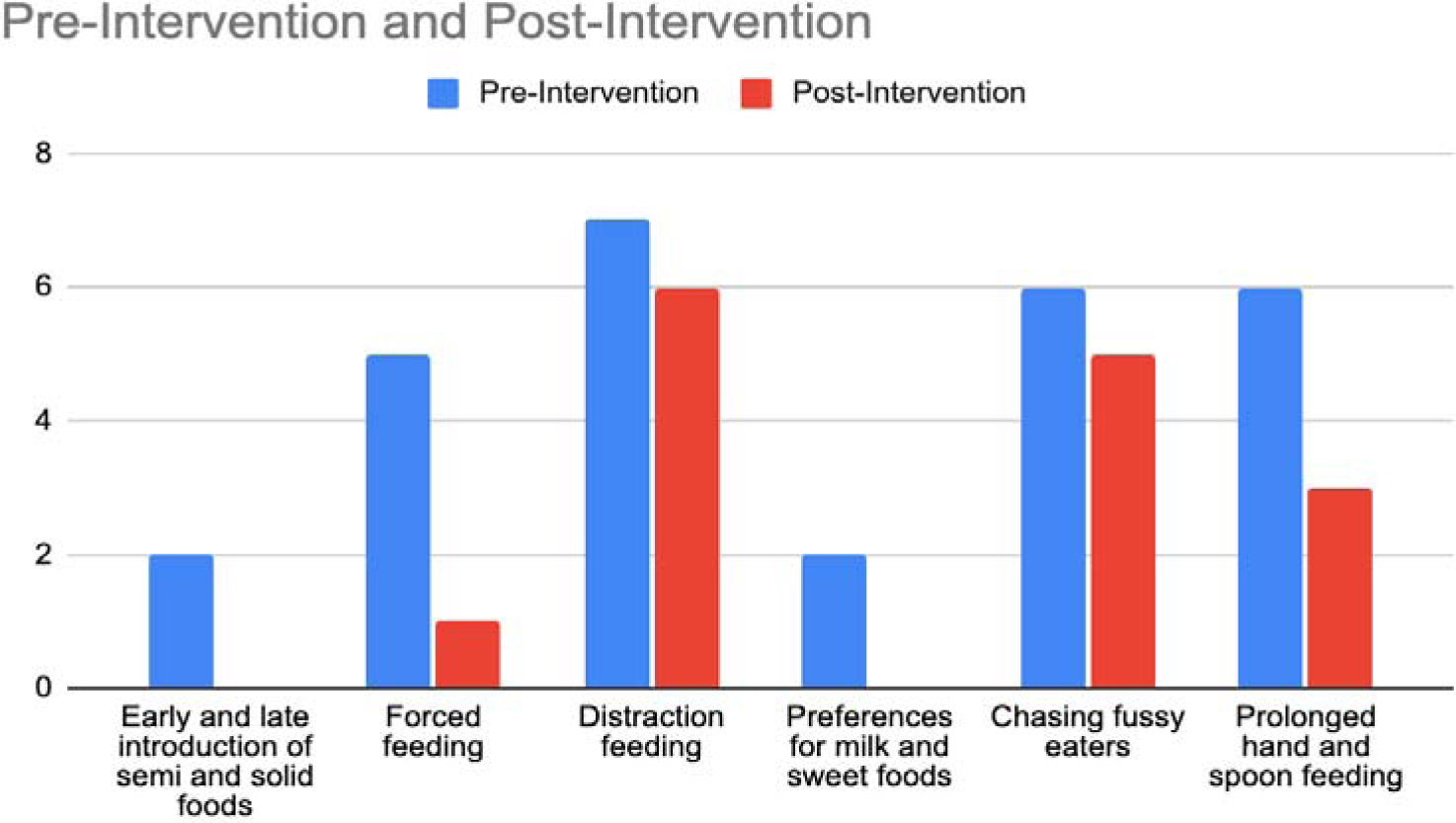
Comparison of feeding styles according to behavioural themes both before and after the intervention

### Outcomes measures for the definitive trial

#### Child Feeding Behaviour Questionnaire

Our study, initially designed to compare infant feeding behaviours, was limited to intervention arm data due to the absence of filled control arm questionnaires. Eight domains at different post-intervention time points were analysed. Notably, ‘food responsiveness’ and ‘food enjoyment’ increased from 29% to 72% and 61% respectively, while ‘emotional overeating’ decreased from 15% to 5%. Other improvements were observed in ‘emotional under-eating’, ‘slowness’, ‘food fussiness’, and ‘responsiveness to food satiety and drinks’. However, response rates declined from 65 to 15 at the 6-month follow-up.

#### Parental Feeding Style

Emotional feeding decreased from 20% to 15% and further to 11% at the 6-month follow-up. Instrumental feeding dropped from 65% to 49% and then to 38% at 6 months. ‘Encouragement to eat’ decreased from 50% to 45%, maintaining this level at the 6-month follow-up. ‘Control overeating’ increased from 40% to 49% and then reduced to 35% at the 6-month follow-up.

#### Feeding Videos

ELAN Linguistic Annotator software analysed feeding videos; coding gestures linked to associated vocabulary. The feeding styles of mothers during the first month were compared with the seventh session of the intervention. In total, 27 feeding videos were collected. No force-feeding was observed in face-to-face sessions, but it occurred in six videos from online sessions. Online mothers were more likely to use toys and media for distraction (12 videos vs. 1 video). Videos before the intervention showed a tendency to overfeed, including force-feeding, distraction feeding, feeding to fill the belly, and prolonged parent-led feeding style. Follow-up videos showed a reduction in force-feeding (1/7 vs. 3/7) but an increase in using objects or phones for distraction during feeding (6/7 vs. 3/7). Three mothers actively assisted children despite their ability to self-feed, and three children fed themselves using hands rather than utensils.

#### Network Diffusion

The intended assessment of study material distribution via e-redbook downloads encountered obstacles. The e-redbook application was unprepared for selective material sharing, risking exposure to unintended recipients. Complications in app registration and inadequate responses to the dissemination survey further exacerbated these issues.

#### Four-Day Food Diary

Response to the food diary survey was exceptionally low due to its detailed nature, perceived burden, and challenges in literacy levels. The CEQ and PFQ surveys, simpler with CF support, were more successful. Low English literacy within the population hindered participants from completing the diary, highlighting the need for user-friendly tools.

#### Equality Impact Assessment (EIA)

EIA systematically assessed the intervention’s impact on equality aspects, considering disability, gender, race, age, sexual orientation, and religion/belief. The intervention was adapted to accommodate diverse needs, respecting differences in ethnicity, language, location, and age. Partners reported an inclusive intervention with no potential discrimination areas. Suggestions included involving fathers and acknowledging the intervention’s specific focus on the SA population.

#### Assessment of BMI Z-scores

ANOVA analysis assessed the intervention’s impact on mean BMI z-scores across the three trial arms and data collection points. Significant differences were noted in baseline weight and BMI z-scores among the arms (see supplemental material 5). At the project’s end, weight and BMI z-scores remained significantly different, with the face-to-face arm showing the highest values. The control arm exhibited a reduction in mean Z-score, and after six months, no significant differences were observed in height, weight, or BMI z-scores among the three arms.

#### Sample Size Calculations

G*Power Software estimates an effect size of 0.457 for BMI Z-scores at the project’s end. To achieve this effect size with a 0.05 alpha error and 80% power in a three-group ANOVA, aiming for a critical F value of 10.66, the total estimated sample size is 51. Planning for a 20% loss to follow-up, each arm is targeted to enrol 21 participants.

## Discussion

This pilot RCT evaluated a PLA intervention in HICs, executing its four phases: problem identification, solution identification, solution implementation, and evaluation. Despite pandemic-induced challenges, successful intervention delivery was achieved through real-time mitigation strategies. The collected data met the Go/Stop criteria, deeming the intervention feasible and acceptable. Participant satisfaction underscored the importance of relationships with CRs and CFs. The study supports advancing to a definitive trial with protocol modifications, guided by rigorous evaluation and adherence to Go/Stop criteria.

### Outcome Measures

The study affirms the feasibility of child anthropometric data collection, hinting a positive potential impact on the BMI z-scores in the intervention group. However, the 6-month trial may not fully capture significant changes, underscoring the need for extended follow-ups. Despite short-term limited changes, the intervention’s influence is anticipated to permeate participant networks, engendering enduring effects. The shifting health system, reallocating public health duties to local authorities, offers prospects for community-centric healthcare, fostering healthier societies. Baseline, post-PLA, and six-month questionnaire data for CEBQ, PFQS, and EIA were collected. Covid-induced delays necessitated modifications to the original one-year follow-up plan.

### Strengths

The seamless data collection in TH, a result of the council and general practitioner (GP) lead’s collaboration, emphasizes the significance of teamwork in successful initiatives. The superior relationship building in the face-to-face arm, compared to the online platform, profoundly impacted participants. Fathers had expressed interest in participating in sessions to educate themselves. The recruitment process was robust in both boroughs due to collaborative efforts and the engagement of advocacy interpreters in TH to facilitate consent recordings was noteworthy. Lastly, Women and Children First’s (WCF) bespoke training enhanced CFs’ guidance and motivation during PLA sessions, underscoring the importance of continuous learning and capacity building for project progression.

### Challenges

RCT design rigidity led to dropouts, necessitating a flexible, participant-preferred delivery mode such as a hybrid model to improve retention. Data collection faced challenges like low-response rates and recording especially in the control arm. In TH, walk-in clinics facilitated measurements, while in NH, appointments with health visitors were required. Low response rates led to HVs directly calling for improved outcomes in NH. Secondary outcome measures, gathered through comprehensive online surveys, encountered low response rates. Difficulties in live session translations and the ineffectiveness of paper forms highlighted the necessity of technological proficiency for online interventions.

The enhancement of community intervention engagement requires a holistic approach, considering factors such as environmental and sociopolitical influences like COVID-19 affecting recruitment from May to September 2022. Careful logistical planning and demographic and cultural considerations are crucial for acceptable interventions and efficient data collection tool design.

## Conclusion

This research sought to address the research question: What is the most acceptable, feasible and robust trial protocol to address the question “*Can a co-adapted participatory learning and action (PLA) cycle targeted at South Asian community members optimise infant feeding, care and dental hygiene practices?*” Based on the trial data collected and our Go/Stop criteria the intervention is deemed feasible and acceptable to community members.

In essence, community-oriented healthcare, underpinned by a strong evidence base and clear narrative, can foster a healthier society. Community-based interventions using PLA approaches have shown potential in promoting health equity in HICs. NEON’s PLA cycle resulted in promising behavioural changes at individual and familial levels. It yielded positive results, underscoring the need for full-cycle completion for comprehensive impact assessment. Future research should utilise the PLA methodology to explore its effectiveness in addressing public health issues in HICs.

## Footnotes

## Supporting information

Supplemental Material 5. Comparison of mean weight and BMI z-scores among the participants of face-to-face PLA, online, and the control groups

Supplemental Material 4. Direct observation score from each session

Supplemental Material 3. Attendance Rate for the PLA sessions

Supplemental Material 2. Breakdown of participants recruited by ethnicity at the start of the trial and at the end of the trial

Supplemental Material 1. Overview schematic detailing number of wards & PLA groups by East London boroughs

## Data Availability

All data produced in the present work are contained in the manuscript

## Acknowledgements

The authors would like to thank the South Asian community facilitators in the NEON Intervention and community members of the London Boroughs of Tower Hamlet and Newham for their important contribution and engagement to this research project. Furthermore, we would like to acknowledge the contribution of the NEON Core Team, Steering Team and all health experts who contributed to this study and validating the intervention and NEON toolkit. We want to thank the Women & Children First Charity and First Steps Nutrition Trust for their valuable contributions and guidance throughout the study. Steering team members had an opportunity to critically review results and contribute to the process of finalising this paper. The authors would like to thank the National Institute of Health Research (NIHR) Academy and the NIHR Collaboration for Leadership in Applied Health Research and Care North Thames for funding the NEON study. This work is also supported by the NIHR GOSH BRC. The views expressed are those of the author(s) and not necessarily those of the NHS, the NIHR or the Department of Health.

## Author’s Contributions

Contributors ML, LM, MH, NB, CL and PP formed the core team of the study and delivered the research methodology. PP carried out all the activities in the intervention delivery and wrote and devised the paper along with TK, SC, DD, KS and SF. LM, ML, MH, NB, CL, JW, OO, JG, KW-M, CI, MA, and PK validated the study and revised the manuscript critically for important intellectual content. PP, TK, SC and DD contributed to the manuscript writing, and prepared it for submission. PP, ML, MH and LM had primary responsibility for the final content. All authors read and contributed to reviewing the study data, the designing of the manuscript, and the approval of the final manuscript.

## Collaborators

In addition to the authors, members of the NEON steering team consist of Prof Atul Singhal, Prof Mitch Blair, Kelley Webb Martin, Carol Irish, Dr Mfon Archibong, Joanna Drazdzewska, Dr Sonia Ahmed, Amelie Gonguet, Gary Wooten, Dr Ian Warwick, Vaikuntanath Kakarla, Phoebe Kalungi, Jenny Gilmore, Prof Richard Watt, Prof Audrey Prost, Dr Edward Fottrell, Ashlee Teakle, Prof Oyinlola Oyebode, Keri McCrickerd, Dr Rana Conway, Professor Lisa Dikomitis, Mari Toomse-Smith, Scott Elliot, Julia Thomas, Aeilish Geldenhuys, Chris Gedge, Kristin Bash, Dr Dianna Smith, Kate Questa, Dr Megan Blake, Gary Tse, Dr Queenie LAW Pui Sze, Gavin Talbot, Dr Chiong Yee Keow, Angela Trude, Lindsay Forbes, Nazanin Zand, Dr Clare Llewellyn, Lakmini Shah, Yeqing Zhang and Natasha Chug.

Steering team members had an opportunity to critically review results and contribute to the process of finalising this paper.

## Patient and public involvement

Patients and/or the public were involved in the design, or conduct, or reporting, or dissemination plans of this research. Refer to the Methods section for further details.

## Disclaimer

The views expressed in the publication are those of the author(s) and not necessarily those of the sponsor (UCL), funder (NIHR), study partners (Tower Hamlets GP Care Group, London Borough of Newham Council).

## Participant consent for publication

Participant information sheets and consent forms were provided to community members and those expressing interest. Community participants agreed to participate and gave audio/video consent prior to their participation in the workshops. Verbal consent was witnessed and formally recorded. All participants were informed of their right to freely withdraw from the study at any time. Confidentiality of personal data was ensured through the use of anonymisation techniques as stated in the Data Protection Act (1998) and in line with the General Data Protection Regulation (2018). All participant data is anonymised and stored on an encrypted password protected computer. Data can only be accessed by the authorised research personnel.

## Ethics approval

This study has obtained ethical approval from UCL Research Ethics Committee [Ethics ID 17269/001], Sponsor reference number: 142600, Funding Reference: NIHR300020 and IRAS number: 296259, Ref: 21/SW/0142. Study protocols and relevant documents were reviewed by UCL Research Ethics Committee, NHS Health Research Authority (HRA) and study partners involved in establishing data sharing agreement for linking participants’ routine data (Tower Hamlets GP Care Group, London Borough of Newham Council).

## Provenance and Peer Review

Not commissioned; peer reviewed for ethical and funding approval prior to submission.

## Data Sharing Statement

The data supporting the findings of this study is available upon reasonable request from the corresponding author.

## Supplementary Materials

**Supplemental Material 1.**
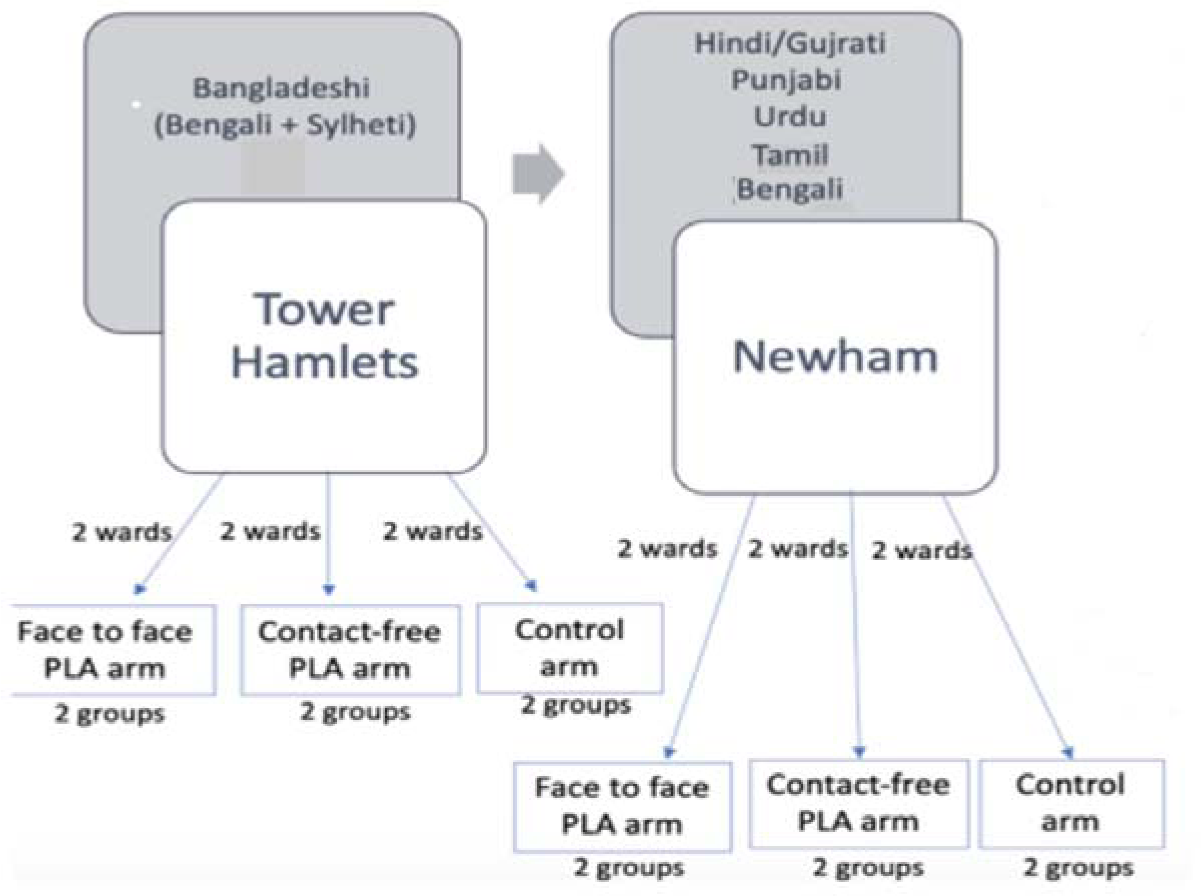
Overview schematic detailing number of wards & PLA groups by East London boroughs

**Supplemental Material 2.**
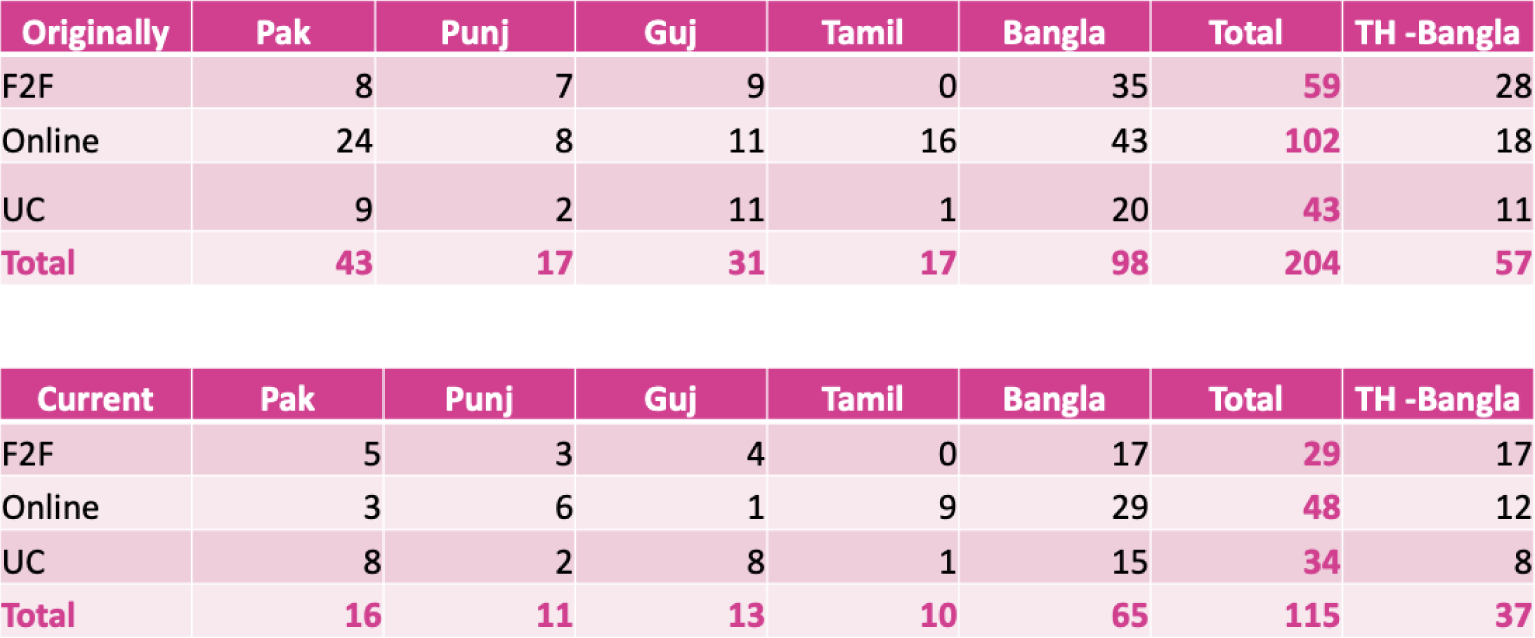
Breakdown of participants recruited by ethnicity at the start of the trial and at the end of the trial

**Supplemental Material 3.**
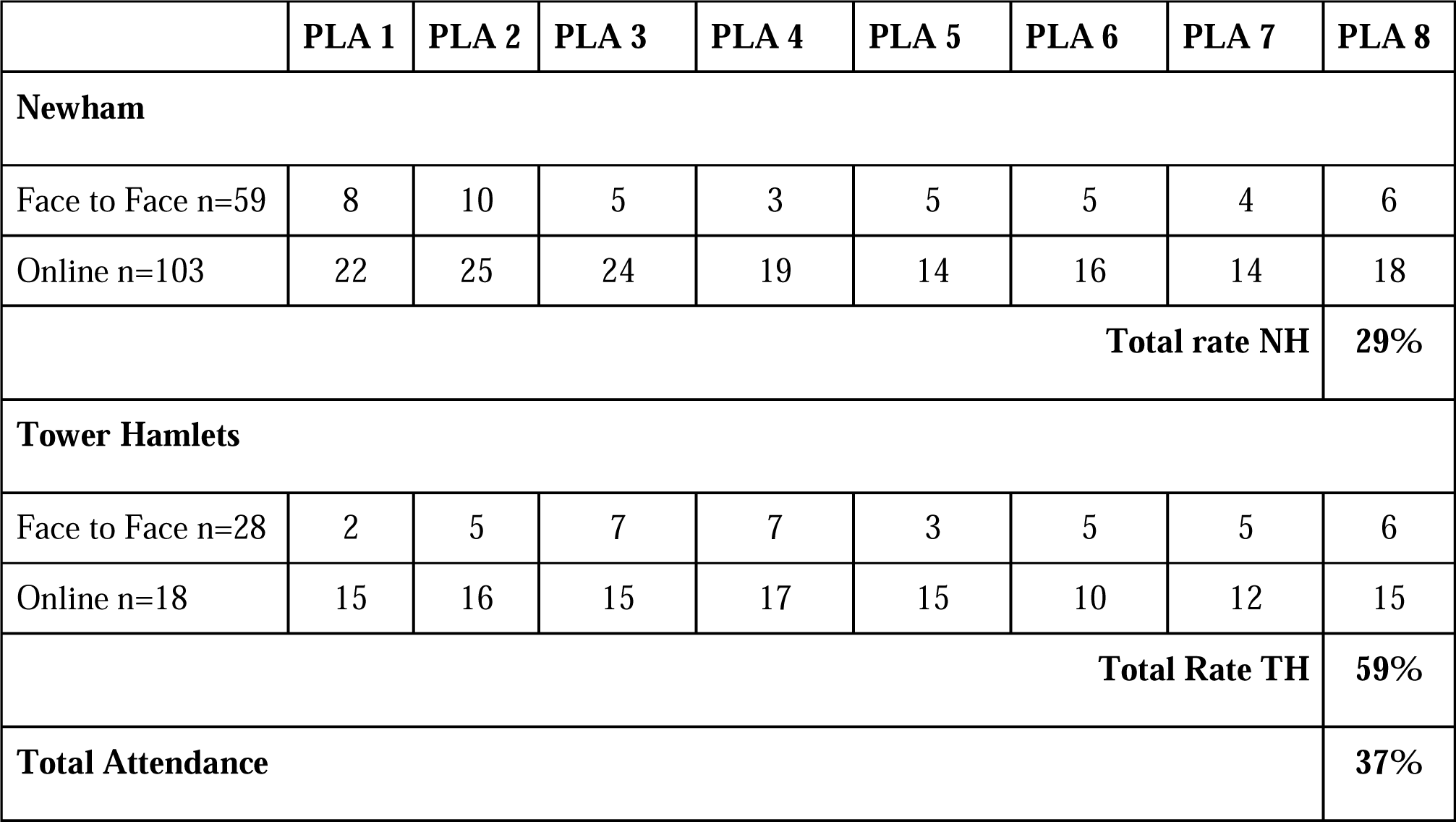
Attendance Rate for the PLA sessions

**Supplemental Material 4.**
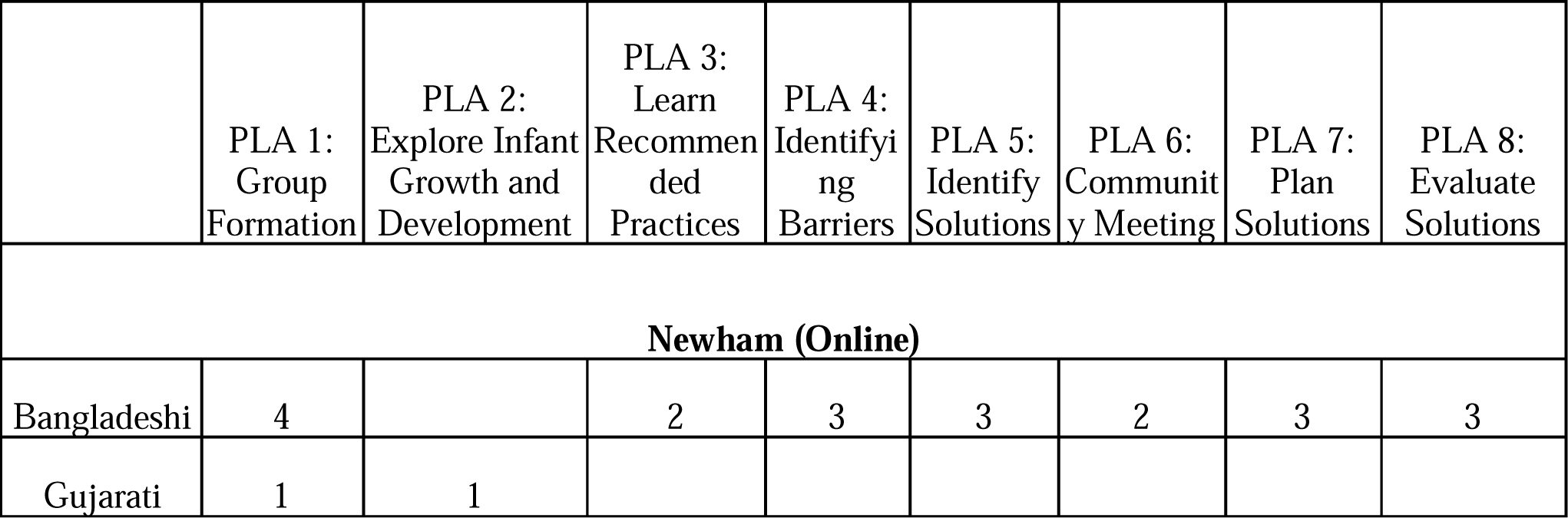
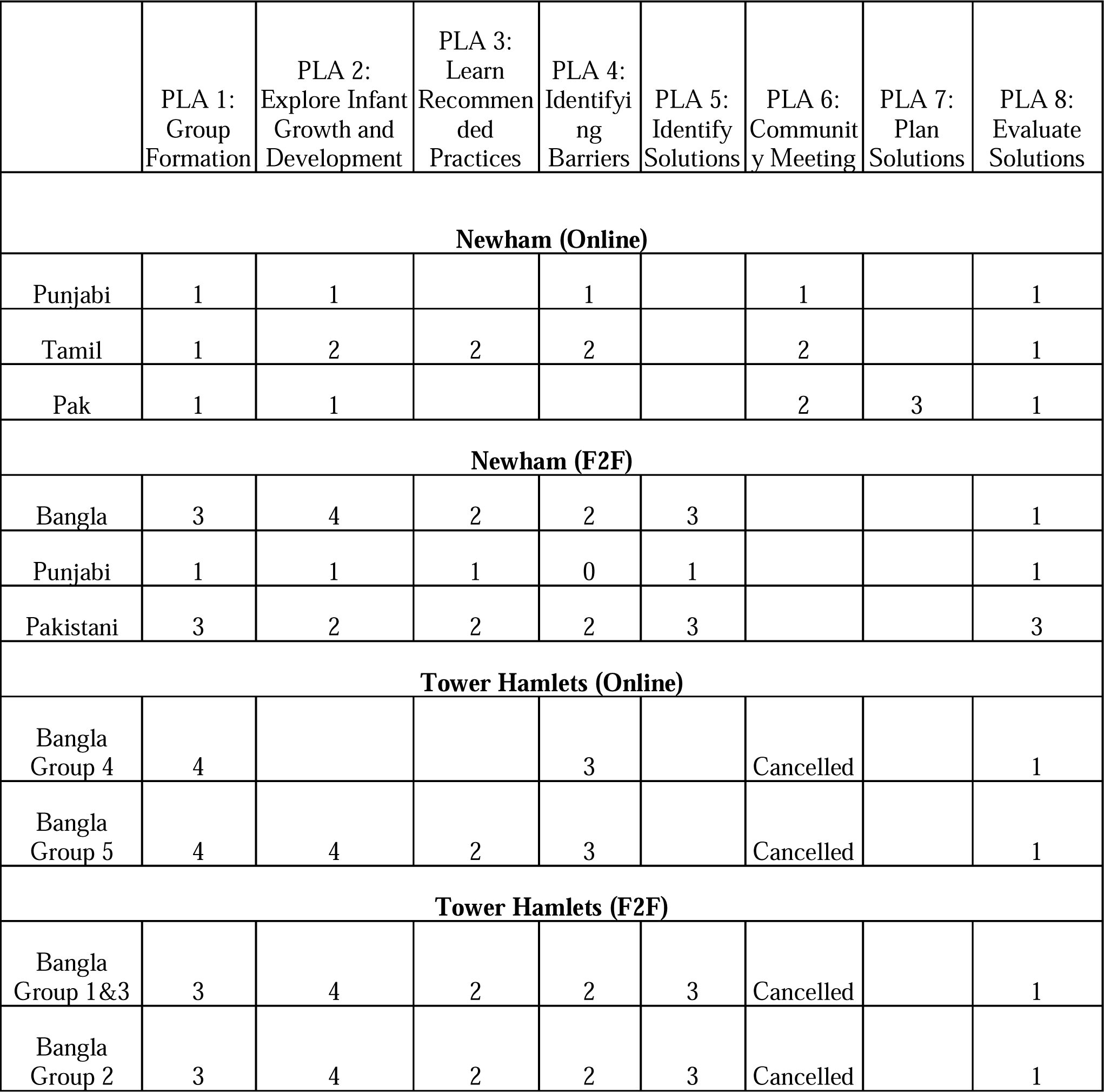
Direct observation score from each session

**Supplemental Material 5.**
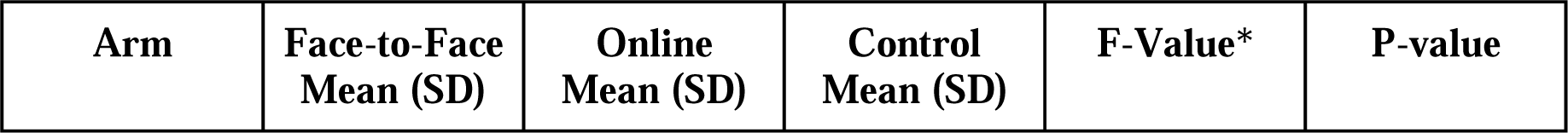
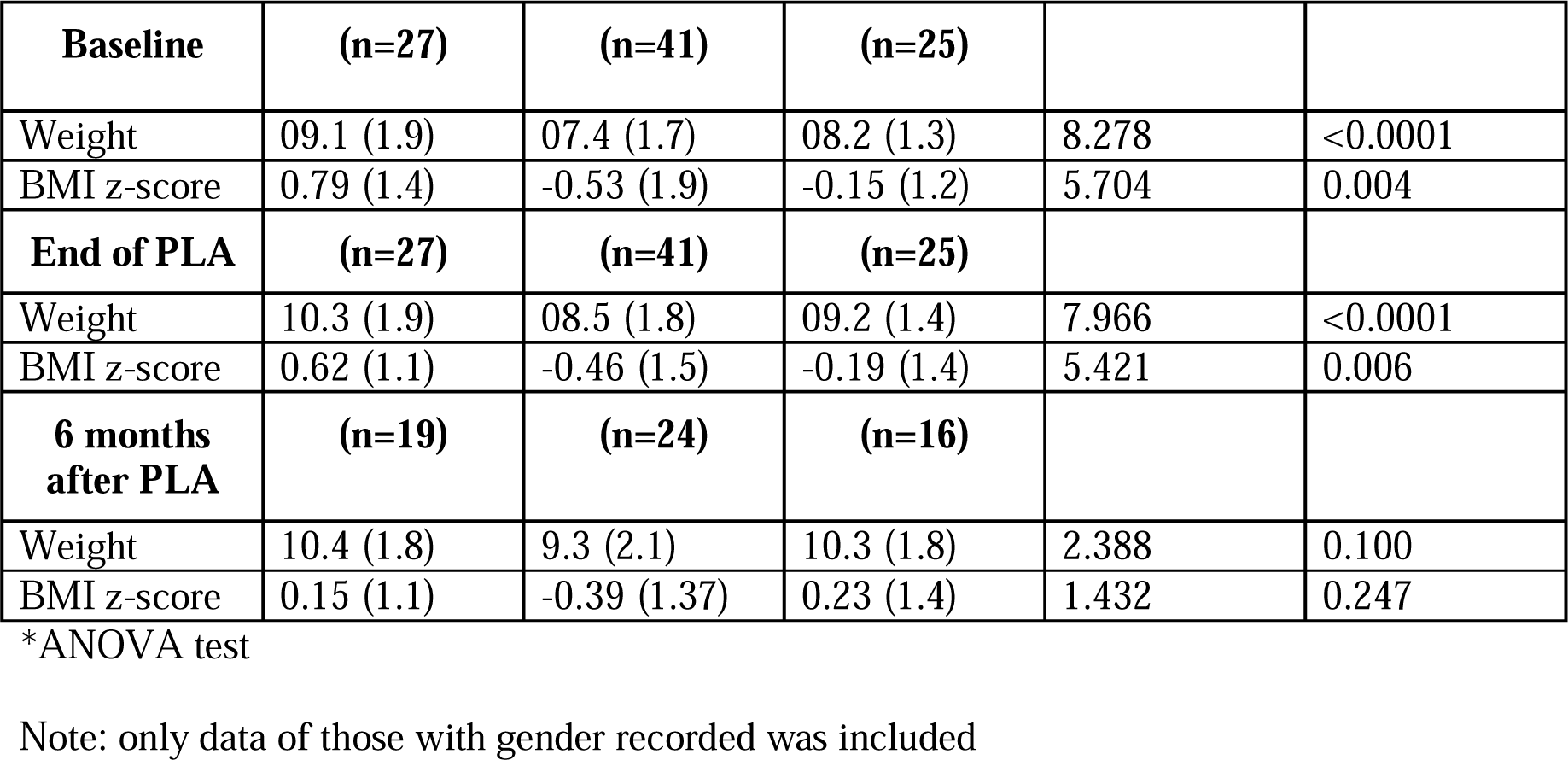
Comparison of mean weight and BMI z-scores among the participants of face-to-face PLA, online, and the control groups

