## Supplemental Material 5. Comparison of mean weight and BMI z-scores among the participants of face-to-face PLA, online, and the control groups for "Pilot Randomised Controlled Trial - Nurture Early for Optimal Nutrition (NEON) Study: Community facilitator-led participatory learning and action (PLA) women’s groups to improve infant feeding, care and dental hygiene practices in South Asian infants aged < 2 years in East London"

| **Arm** | **Face-to-Face**  **Mean (SD)** | **Online**  **Mean (SD)** | **Control**  **Mean (SD)** | **F-Value*** | **P-value** |
| --- | --- | --- | --- | --- | --- |
| **Baseline** | **(n=27)** | **(n=41)** | **(n=25)** |  |  |
| Weight | 09.1 (1.9) | 07.4 (1.7) | 08.2 (1.3) | 8.278 | <0.0001 |
| BMI z-score | 0.79 (1.4) | -0.53 (1.9) | -0.15 (1.2) | 5.704 | 0.004 |
| **End of PLA** | **(n=27)** | **(n=41)** | **(n=25)** |  |  |
| Weight | 10.3 (1.9) | 08.5 (1.8) | 09.2 (1.4) | 7.966 | <0.0001 |
| BMI z-score | 0.62 (1.1) | -0.46 (1.5) | -0.19 (1.4) | 5.421 | 0.006 |
| **6 months after PLA** | **(n=19)** | **(n=24)** | **(n=16)** |  |  |
| Weight | 10.4 (1.8) | 9.3 (2.1) | 10.3 (1.8) | 2.388 | 0.100 |
| BMI z-score | 0.15 (1.1) | -0.39 (1.37) | 0.23 (1.4) | 1.432 | 0.247 |

*ANOVA test

Note: only data of those with gender recorded was included
