## Supplemental Material 4. Direct observation score from each session for "Pilot Randomised Controlled Trial - Nurture Early for Optimal Nutrition (NEON) Study: Community facilitator-led participatory learning and action (PLA) women’s groups to improve infant feeding, care and dental hygiene practices in South Asian infants aged < 2 years in East London"

|  | PLA 1: Group Formation | PLA 2: Explore Infant Growth and Development | PLA 3: Learn Recommended Practices | PLA 4: Identifying Barriers | PLA 5: Identify Solutions | PLA 6: Community Meeting | PLA 7: Plan Solutions | PLA 8: Evaluate Solutions |
| --- | --- | --- | --- | --- | --- | --- | --- | --- |
| **Newham (Online)** | | | | | | | | |
| Bangladeshi | 4 |  | 2 | 3 | 3 | 2 | 3 | 3 |
| Gujarati | 1 | 1 |  |  |  |  |  |  |
| Punjabi | 1 | 1 |  | 1 |  | 1 |  | 1 |
| Tamil | 1 | 2 | 2 | 2 |  | 2 |  | 1 |
| Pak | 1 | 1 |  |  |  | 2 | 3 | 1 |
| **Newham (F2F)** | | | | | | | | |
| Bangla | 3 | 4 | 2 | 2 | 3 |  |  | 1 |
| Punjabi | 1 | 1 | 1 | 0 | 1 |  |  | 1 |
| Pakistani | 3 | 2 | 2 | 2 | 3 |  |  | 3 |
| **Tower Hamlets (Online)** | | | | | | | | |
| Bangla Group 4 | 4 |  |  | 3 |  | Cancelled |  | 1 |
| Bangla Group 5 | 4 | 4 | 2 | 3 |  | Cancelled |  | 1 |
| **Tower Hamlets (F2F)** | | | | | | | | |
| Bangla Group 1&3 | 3 | 4 | 2 | 2 | 3 | Cancelled |  | 1 |
| Bangla Group 2 | 3 | 4 | 2 | 2 | 3 | Cancelled |  | 1 |
