## Supplemental Material 3. Attendance Rate for the PLA sessions for "Pilot Randomised Controlled Trial - Nurture Early for Optimal Nutrition (NEON) Study: Community facilitator-led participatory learning and action (PLA) women’s groups to improve infant feeding, care and dental hygiene practices in South Asian infants aged < 2 years in East London"

|  | **PLA 1** | **PLA 2** | **PLA 3** | **PLA 4** | **PLA 5** | **PLA 6** | **PLA 7** | **PLA 8** |
| --- | --- | --- | --- | --- | --- | --- | --- | --- |
| **Newham** | | | | | | | | |
| Face to Face n=59 | 8 | 10 | 5 | 3 | 5 | 5 | 4 | 6 |
| Online n=103 | 22 | 25 | 24 | 19 | 14 | 16 | 14 | 18 |
| **Total rate NH** | | | | | | | | **29%** |
| **Tower Hamlets** | | | | | | | | |
| Face to Face n=28 | 2 | 5 | 7 | 7 | 3 | 5 | 5 | 6 |
| Online n=18 | 15 | 16 | 15 | 17 | 15 | 10 | 12 | 15 |
| **Total Rate TH** | | | | | | | | **59%** |
| **Total Attendance** | | | | | | | | **37%** |
