## Supplemental Material 1. Overview schematic detailing number of wards & PLA groups by East London boroughs for "Pilot Randomised Controlled Trial - Nurture Early for Optimal Nutrition (NEON) Study: Community facilitator-led participatory learning and action (PLA) women’s groups to improve infant feeding, care and dental hygiene practices in South Asian infants aged < 2 years in East London"

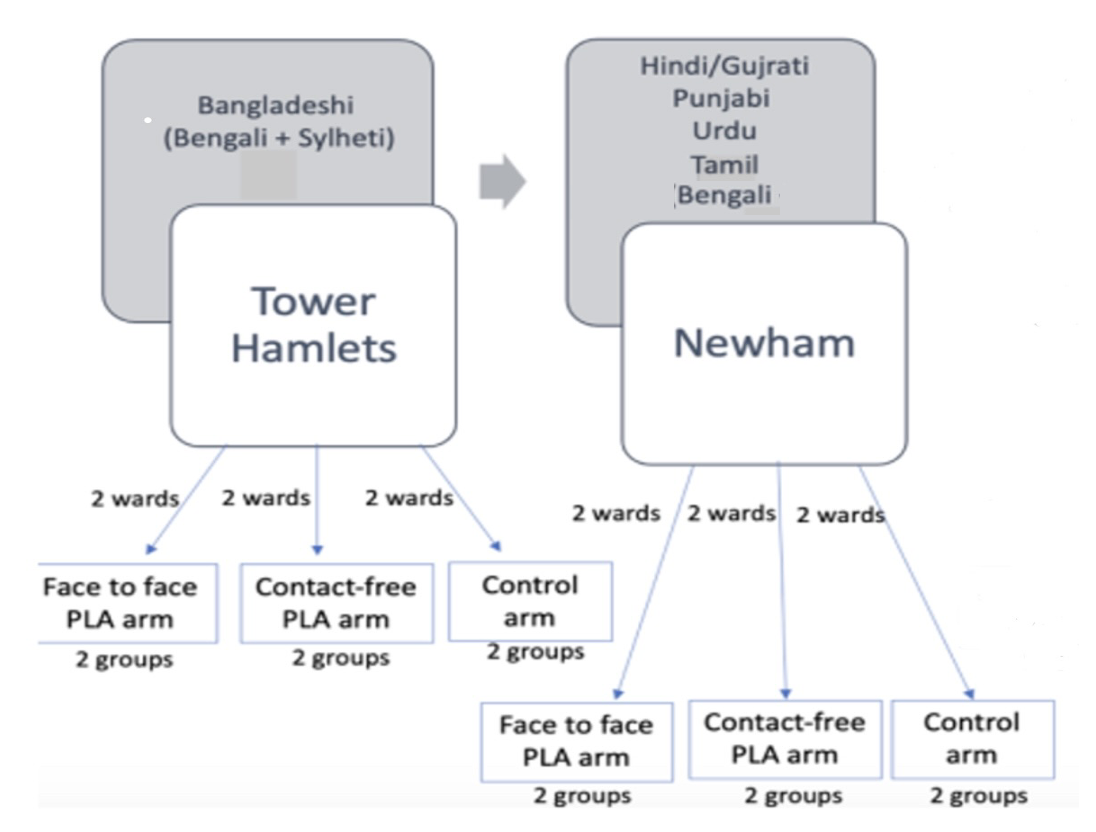
